# Use and impact of virtual primary care on quality and safety: the public’s perspectives during the COVID-19 pandemic

**DOI:** 10.1101/2021.10.19.21265193

**Authors:** Ana Luisa Neves, Jackie van Dael, Niki O’Brien, Kelsey Flott, Saira Ghafur, Ara Darzi, Erik Mayer

## Abstract

**Background:** With the onset of COVID-19, primary care has swiftly transitioned from face-to-face to virtual care, yet it remains largely unknown how this has impacted on the quality and safety of care.

**Aim:** To evaluate patient use of virtual primary care models during COVID-19 in terms of change in uptake, perceived impact on the quality and safety of care, and willingness of future use.

**Design and setting:** An online cross-sectional survey was administered to the public across the United Kingdom, Sweden, Italy and Germany.

**Methods:** McNemar tests were conducted to test pre- and post pandemic differences in uptake for each technology. One-way analysis of variance was conducted to examine patient experience ratings and perceived impacts on healthcare quality and safety across demographic characteristics.

**Results:** Respondents (N=6,326) reported an increased use of telephone consultations (+6.3%, P<.001), patient-initiated services (+1.5%, n=98, p<0.001), video consultations (+1.4%, P<.001), remote triage (+1.3, p<0.001), and secure messaging systems (+0.9%, P=.019). Experience rates using virtual care technologies were higher for men (2.39±0.96 vs 2.29±0.92, P<.001), those with higher literacy (2.75±1.02 vs 2.29±0.92, P<.001), and participants from Germany (2.54±0.91, P<.001). Healthcare timeliness and efficiency were the quality dimensions most often reported as being positively impacted by virtual technologies (60.2%, n=2,793 and 55.7%, n=2,401, respectively), followed by effectiveness (46.5%, n=1,802), safety (45.5%, n=1,822), patient-centredness (45.2%, n=45.2) and equity (42.9%, n=1,726). Interest in future use was highest for telephone consultations (55.9%), followed by patient-initiated digital services (56.1%), secure messaging systems (43.4%), online triage (35.1%), video consultations (37.0%), and chat consultations (30.1%), although significant variation was observed between countries and patient characteristics.

**Conclusion:** Future work must examine the drivers and determinants of positive experiences using remote care to co-create a supportive environment that ensures equitable adoption and use across different patient groups. Comparative analysis between countries and health systems offers the opportunity for policymakers to learn from best practices internationally.

## Introduction

The delivery of health and care services has evolved rapidly following the onset of the COVID-19 pandemic. To mitigate the transmission of COVID-19, and respond to the increased burden of care, the pandemic has advanced the use of virtual care technologies in facilitating care for patients globally, disrupting traditional face-to-face care models. Virtual remote care may be delivered via telephone or text, or video and other online tools which require internet connection. In England, based on updated policy guidance, primary care practitioners have adapted their models of care to offer over 85% of consultations remotely via phone or video link, where previously 95% of consultations took place face-to-face.^1^ Similarly, in Italy the Digital Solidarity initiative launched by the Minister for Technological Innovation and Digitization, offers online public or specialist services in response to the pandemic, including health services such as “Video Visita” (Video Visit), which offers patients free video consultations with family doctors and paediatricians signed up to the service.^2^

While the use of virtual care accelerated rapidly in response to the COVID-19 pandemic, the digitisation of healthcare has been advancing over decades,^3^ as has the utilisation of digital technology in healthcare delivery as a policy ambition at the global level.^4^ For example, in the UK, the NHS Five Year Forward Review, for example, outlined plans for England to utilise digital technologies to offer patients more convenience in accessing care.^5^ However, uptake at scale prior to the pandemic was slow and challenging.

While part of an emergency response, the rapid adoption of virtual care solutions in General Practice will likely have long-term effects on how services are delivered, there is little evidence on its impact on the quality, safety, and equity of care.^6^ There is also sparse evidence on the lessons learned through the experience of delivering large-scale virtual services during the pandemic, which could be utilised to lay the foundations for high-quality virtual care delivery in the post-pandemic future. To facilitate safe, sustainable adoption of virtual models for health and care delivery, it is critical to listen to the perspectives of patients as end-users. Targeted public research must seek to understand their perspectives on the impact of the care received and the risk of widening health inequities between different groups of patients.

This study aimed to evaluate the impact of the adoption of virtual technologies by patients for primary care access during the COVID-19 pandemic. The primary aims were to evaluate the use of virtual care solutions by patients during the COVID-19 crisis, explore different rates of adoption and perceptions across different patient groups, to understand the perceived impact on quality and safety of care during the crisis, and to examine how they would like to continue using virtual primary care, in the post-pandemic future.

## Methods

### Study design

A cross-sectional online survey was performed, adhering to the STrengthening the Reporting of OBservational studies in Epidemiology (STROBE) guideline for cross-sectional studies.^7^

### Participants

Eligible study participants were adults aged over 18 years who were able to speak, read and write in their resident country’s language. Recruitment was carried out with YouGov, an international research data and analytics group. Participants were recruited by YouGov from a host of different sources, including via standard advertising, and strategic partnerships with a broad range of websites. Stratified sampling was used to recruit responses from four countries (Germany, Italy, Sweden, United Kingdom) and, to ensure nationally representative samples, YouGov drew a sub-sample representative of national adults in terms of age, gender, social class, and education level. Data collection took place in December 2020. Ethical approval was granted by Imperial College London’s Ethics Research Committee (Approval number: 20IC5956).

### Survey development

The questions of the survey were developed based on a scoping review and expert consultation. A draft was piloted with GPs/FDs across 20 countries, and feedback incorporated in the final version. The final survey included four sections with a total of 13 items.

The first section examined patients’ characteristics (i.e., gender, age, ethnicity, overall health status, digital health literacy). Digital health literacy was assessed using the eHealth Literacy Scale (eHEALS), a validated tool that reflects the individuals’ own perception of their knowledge and skills at using eHealth information.^8^ The validity and reliability of eHEALS has been demonstrated in several health conditions,^9,10^ and has been translated into various languages.^11,12^ As in previous studies, the eHEALS score was categorised into high eHealth (eHEALS≥26 out of 40) and low eHealth literacy levels (eHEALS<26).^9,11,12^

The second section explored patients’ use of virtual care care before and during the pandemic, including the use of telephone consultations, video consultations, chat consultations, remote triage, patient-initiated digital services (i.e., scheduling, prescription renewal, test requests, sick leave), or secure messaging systems (i.e., to receive test results or other information). Patients’ experiences of these models of care were assessed using a free-text question “In general, how was your overall experience using these technologies?” and a five-point Likert scale (“1 – very bad” to “5 – very good”).

The third section assessed patients’ perceptions on the impact of these technologies on several dimensions of quality and safety of care, as defined by the Institute of Medicine, including: patient-centredness, effectiveness, efficiency, safety, timeliness and equity.^13^

The fourth section explored respondents’ willingness of future use of each investigated virtual care technology.

The online survey was hosted on the Qualtrics platform. A copy of the Qualtrics survey is provided as **Supplementary File 1**.

### Data analysis

Quantitative data were analysed using Statistical Package for the Social Sciences (SPSS) v.27. Descriptive statistics were calculated, including absolute (n) and relative frequencies (%) for categorical variables, and mean (μ) and standard deviation (SD) for continuous variables. McNemar tests were used to examine differences in adoption of each respective technology across countries before versus during the COVID-19 pandemic. One-way analysis of variance (ANOVA) was used to test group-based differences in rated experience of using virtual technologies. Chi-square tests of independence were generated to test associations between demographic characteristics and perceived impacts of virtual care technologies on the quality and safety of care, as well as willingness to use virtual care technologies in the future. The significance level for all statistical tests were set at p-value (P) <0.05, two-tailed.

## Results

### Participants’ characteristics

A total of 6,325 participants (50.0% women) replied to the survey. A detailed description of the characteristics of the sample can be found in **Table 1**, including a breakdown per country.

**Table 1.** Characteristics of the respondent population (n=6,326), by country.

|  | UK<br>(n=2,019) | Germany<br>(n=2,161) | Italy<br>(n=1,131) | Sweden<br>(n=1,015) | Total<br>(n=6,326) |
| --- | --- | --- | --- | --- | --- |
| **Age, n (%)** |  |  |  |  |  |
| 18-24 | 214 (10.6) | 196 (9.1) | 92 (8.1) | 84 (8.3) | 586 (9.3) |
| 25-34 | 326 (16.1) | 330 (15.3) | 152 (13.4) | 205 (20.2) | 1,013 (16.0) |
| 35-44 | 320 (15.8) | 314 (14.5) | 193 (17.0) | 143 (14.1) | 970 (15.3) |
| 45-54 | 327 (16.2) | 388 (18.0) | 221 (19.5) | 155 (15.3) | 1,090 (17.2) |
| 55+ | 831 (41.2) | 933 (43.2) | 474 (41.9) | 428 (42.2) | 2,668 (42.2) |
| **Gender, n (%)** |  |  |  |  |  |
| Female | 1,038 (51.4) | 1,103 (51.0) | 593 (52.4) | 507 (50.0) | 3,241 (51.2) |
| Male | 981 (48.6) | 1,058 (49.0) | 538 (47.6) | 508 (50.0) | 3,085 (48.8) |
| **Ethnicity, n (%)** |  |  |  |  |  |
| Asian | 59 (2.9) | 0 (0.0) | 4 (0.4) | 21 (2.1) | 84 (1.3) |
| Black / African /<br>Caribbean | 21 (1.0) | 0 (0.0) | 5 (0.4) | 6 (0.6) | 32 (0.5) |
| Mixed / Multiple<br>Ethnic Groups | 48 (2.4) | 0 (0.0) | 65 (5.7) | 45 (4.4) | 158 (2.5) |
| Other Ethnic Group | 25 (1.2) | 0 (0.0) | 19 (1.7) | 11 (1.1) | 55 (0.9) |
| White | 1,866 (92.4) | 0 (0.0) | 1,019 (90.0) | 914 (90.0) | 3,798 (60.0) |
| Prefer not to say | 0 (0.0) | 2,161 (100.0) | 20 (1.8) | 18 (1.8) | 2199 (34.8) |
| **eHealth Literacy** | 17.8 (6.4) | 20.0 (7.0) | 20.1 (6.5) | 17.0 (6.7) | 18.8 (6.8) |
| (eHEALS), $\mu$ (SD) | | | | | |

No significant differences were found in the composition of the national samples regarding gender (χ^2^=1.376, P=0.711). The proportion of respondents aged 25–34 years was slightly higher in the UK (16.1%) and Sweden (20.2%) compared to Germany (15.3%) and Italy (13.4%), (χ^2^=36.277, p<.001).

The proportion of white respondents was higher in the UK (92.4%) than in Sweden (90.0%) and Italy (90.0%) P<.001). The UK had a lower proportion of Multiple Ethnic respondents compared to Sweden and Italy (2.4% vs. 4.4% and 5.7%) (P<.001). In Germany, it was not possible to collect ethnicity data due to GDPR restrictions.

Digital literacy was lowest in Sweden (17.03±6.70), compared to the UK (17.82±6.39) (P=0.009), Germany (20.02±6.98) (P<.001), and Italy (20.06±6.51) (P<.001).

### Use of virtual primary care before and during the COVID-19 pandemic

Before the COVID-19 pandemic, highest usage was observed for remote patient-initiated services (19.4%, n=1,228) and secure messaging systems (10.0%, n=632). Use of telephone, video, chat consultations and remote triage were 24.9% (n=1,572), 5.4% (n=341) 6.7% (n=425), and 6.2% (n=392), respectively. During the COVID-19 pandemic, significant increases were observed for telephone consultations (+6.3%, P<.001), patient-initiated services (+1.5%, n=98, P<.001), video consultations (+1.4%, P<.001), remote triage (+1.3, p<.001), and secure messaging systems (+0.9%, P=0.019) **Figure 1A)**.

**Figure 1:**
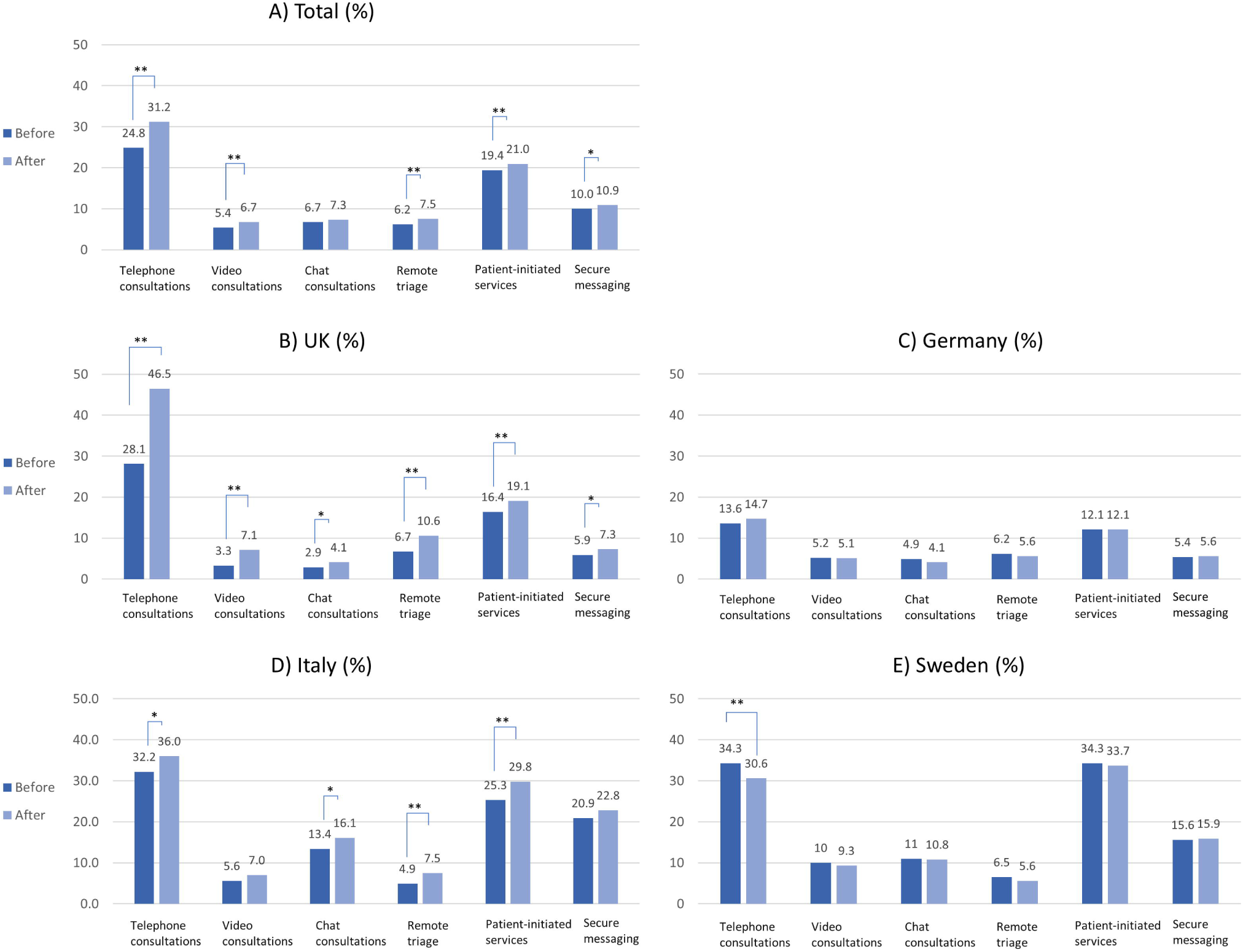
Percentage of usage of each respective technology before and after COVID-19, by country (in%) *Footnote*. *p < .05; **p < .001

In the UK, there was a significant increase for all virtual care technologies, including telephone consultations (+18.4%, p<0.001), remote triage (+3.9%, P<.001), video consultations (+3.8%, P<.001), patient-initiated services (+2.7%, P<.001), secure messaging (+1.4%, P=0.024), and chat consultations (+1.2%, P=0.013) **(Figure 1B)**. In Germany, there was no significant variation in any of the virtual care technologies **(Figure 1C)**. In Italy, there was a significant increase in patient-initiated services (4.5%, P<.001), telephone consultations (3.8%, P=0.004), chat consultations (2.7%, P=0.010), and remote triage (2.6%, P<0.001) **(Figure 1D)**, but not in secure messaging services or video consultations (1.9%, P=0.123 and 1.4%, P=0.072, respectively). In Sweden, telephone consultations decreased significantly (−3.7%, P=0.007), and there was no significant difference in the use of other technologies (**Figure 1E)**.

### Patient experience virtual primary care before and during the COVID-19 pandemic

Of the 3,976 (62.9%) responses to the experience item, the majority reported having had either a good or very good experience (59.8%, n=2,379). Only a small minority reporting a bad or very bad experience (8.4%, n=333). Lower rated experience was observed in women (2.39±0.96 vs 2.29±0.92, P<.001) and those with low literacy respondents (eHEALS≥26) (2.29±0.92 vs 2.75±1.02, P<.001). Patient experience (expressed as the average Likert scale value) significantly varied between countries (P<.001), with participants from Germany reporting a higher average Likert scale value (2.54±0.91) compared to those from the UK (2.24±0.99) (P<.001), Italy (2.28±0.84) (P<.001) and Sweden (2.29±0.96) (P<.001).

### Impact of digital remote care on quality and safety of care during the COVID-19 pandemic

Globally, digital patient-initiated services were most often reported as having a positive impact on the quality of care received across respondents (61.0%, n=2,171), followed by telephone consultations (56.5%, n=2,102), and video consultations (44.1%, n=1,246). Online triage (42.5%, n=1,294), chat consultations (40.3%, n=1,177), and remote monitoring (36.6%, n=968) were least often reported as impacting positively on the quality of care. A breakdown per country for each technology and proportions of missing answers are provided in **Supplementary Tables 1-6**.

Virtual care technologies were most often reported as having positively impacted on timeliness (60.2%, n=2,793) and efficiency (‘avoiding waste’) (55.7%, n=2,401) of care, followed by effectiveness (46.5%, n=1,802), safety (‘avoiding harm’) (45.5%, n=1,822), and patient-centredness (45.2%, n=45.2). Equity was least often reported as having been positively impacted (42.9%, n=1,726). The proportion of missing answers for each domain of quality are reported in **Supplementary Table 7**.

Significant variation was observed in the perceived impact of virtual care technologies on all dimensions of quality between countries (**Figure 2**) (all P<.001). Respondents from Italy were most likely to report a positive impact on each of the six dimensions of care quality. Conversely, respondents in the UK were most likely to report a negative impact on each of the six dimensions of care quality, although all <17.0%.

**Figure 2:**
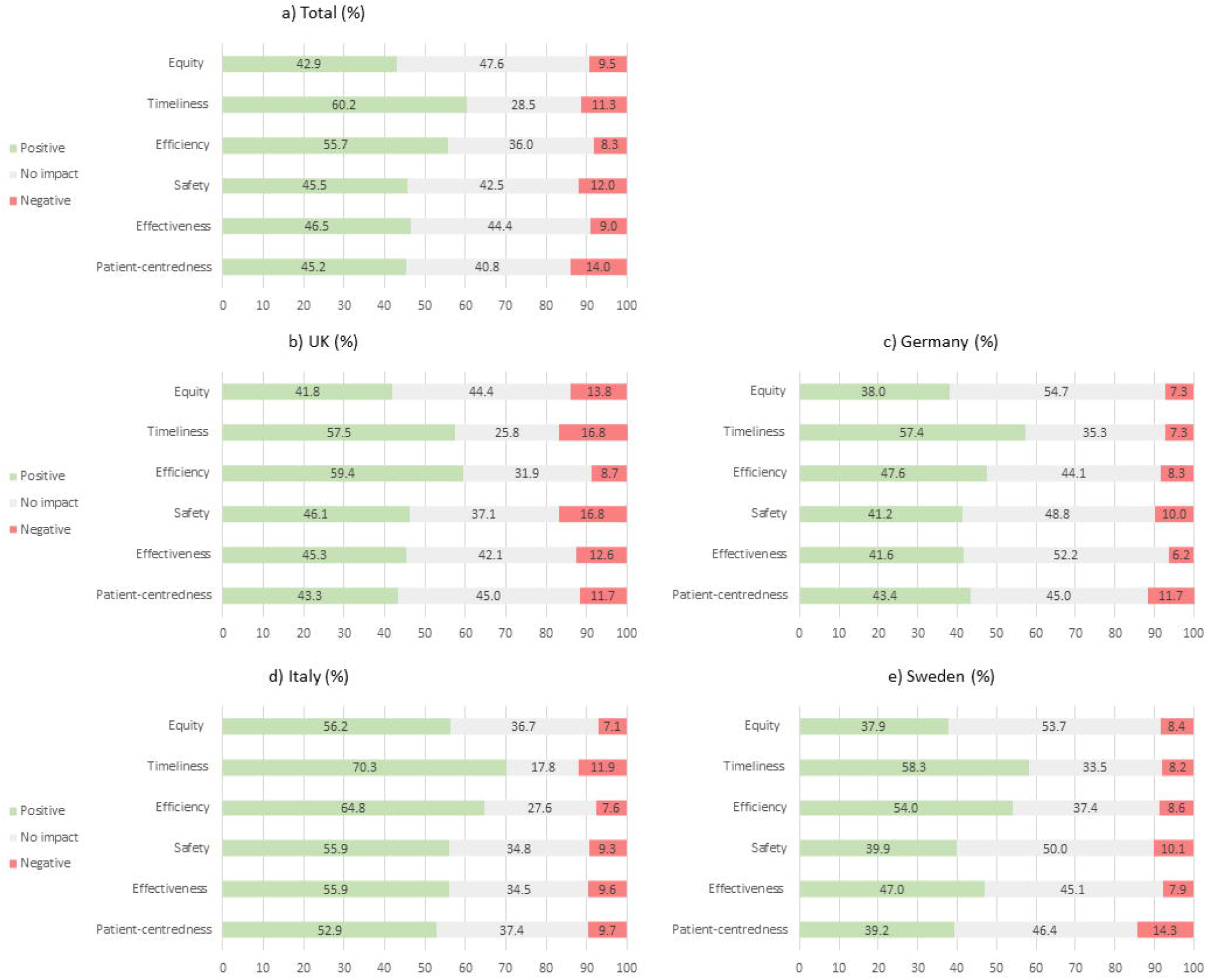
Respondents’ perceptions of how virtual care technologies have impacted on each aspect of care received, by country (in%)

### Willingness to use virtual care technologies after the COVID-19 pandemic

Among those that answered the questions about intended future use, a slight majority of participants reported wanting to continue using telephone consultations (55.9%, n=3,196) and patient-initiated digital services (56.1%, n=3,206) (**Figure 3**). Less than half of the participants expressed willingness to use secure messaging systems (43.4%, n=2,480), online triage (35.1%, n=2,005), video consultations (37.0%, n=2,114) or chat consultations (30.1%, n=1,718) in the future.

**Figure 3:**
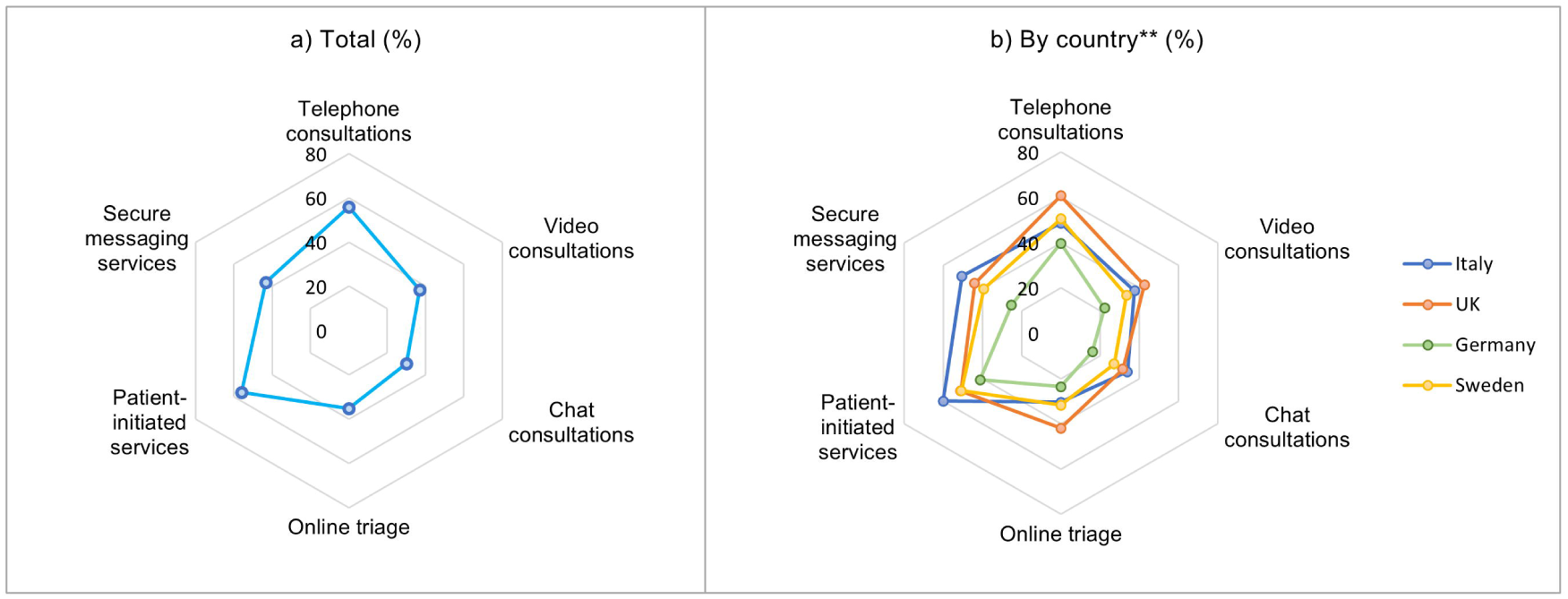
Respondents’ willingness of future use of each respective technology after COVID-19, by country (in %) \*\**Footnote*. Chi-square tests found significant variation across countries for each of six listed technologies (p<.001)

A significant variation was observed between countries (P<.001), with German participants (24.8%, n=477) being least likely to want to use any of the listed virtual care technologies, followed by the UK (14.6%, n=267), Sweden (7.4%, n=50), and Italy (4.8%, n=50). UK participants reported the highest support for future use of telephone consultations (67.1%, n=1,226), video consultations (47.0%, n=858), and online triage (46.3%, n=845). In Sweden, most participants supported the future use of patient-initiated digital services (66.1%, n=606), secure messaging systems (52.2%, n=479), and chat consultations (37.8%, n=347). In Italy, participants were most frequently interested in using patient-initiated digital services (64.4%, n=679), secure messaging systems (54.2%, n=570), and telephone consultations (52.3%, n=550) in the future. German participants showed least frequent willingness in future use of all the technologies evaluated, although nearly half of respondents expressed interest in the future use of telephone consultations (44.5%, n=857) and patient-initiated services (46.3%, n=891).

Willingness to use telephone consultations increased with increasing age; for video, chat consultations and online triage, interest in future use reached a peak at 35-54 years, progressively declining in older groups (all P<.001). Not wanting to use *any* of these remote care solutions was higher in men (14.7 vs 12.6, P=0.017), those with higher digital literacy (24.4 vs 12.0, P<.001), and those in higher age bands (P<.001). Interestingly, participants with higher eHEALS score consistently reported a lower intention to use each one of the remote care technologies evaluated (all P<.001). A detailed overview of these results is provided in **Supplementary Tables 8-12**.

## Discussion

Our findings indicate a stark increase in the use of remote care technologies in Primary Care during the COVID-19 pandemic in the UK and Italy. This is in line with recently published data in the UK, that suggests that, by April 2020, almost half of appointments were reported as taking place remotely.^14^ This sharp rise contrasts with usage in Germany and Sweden, which were found to remain more conservative in the triaging patients to virtual care (despite German participants providing the most positive experience ratings). These differences may reflect the respective health policy landscape in each country, underscoring the potential of lessons to be gained from inter-country comparisons. For example, in contrast to the UK, where commercial products such as Skype, WhatsApp or Facetime are used in urgent cases (if no alternatives available), digital health applications in Germany require approval by the responsible government agencies based on a demonstration of evidence of positive effects in order to be accredited and reimbursed.^15^

Most of our participants reported having had a (very) good experience using remote care. In the UK, Imlach et al (2020) also found that patients reported high satisfaction with virtual care, describing it as a convenient mode of care that allowed them to safely access health care without having to weigh-up the fear of COVID-19 infection against the need to be observed.^16^ Yet, it is important to note that patients’ experiences vary across patients’ characteristics; our results suggest a better experience by those with higher digital health literacy and, in Italy, by some ethnic groups. Timeliness and efficiency were reported as the quality of care dimensions most positively impacted by the introduction of virtual care. Graetz et al (2020) examined patient-initiated primary care visits in a large integrated health care delivery system with over 4 million members and observed that fewer than half of in-person visits were scheduled within one day of making the appointment, compared to over half of video visits, and over two-thirds of telephone visits.^17^ On average, telephone visits were scheduled 50% sooner than office visits. This is in line with findings of previous studies highlighting that remote care can reduce wait times.^18,19^ Previous evidence has also highlighted a potential benefit of remote care regarding the efficacy of physicians, reduced medical costs, and improved the access to health care services,^20^ although evidence is somewhat inconsistent.

Across countries, more than half of the participants wanted to continue using telephone consultations and patient-initiated digital services. Interestingly, participants with higher eHealth literacy consistently reported a lower willingness to use each one of the remote care technologies evaluated – despite reporting higher experience rates. Interest in future use differed significantly by country, with German participants being least likely to want to use any of the listed virtual care technologies, potentially due to the relative novelty of these applications. Our results also suggest that patients with higher eHealth literacy may report a lower willingness to use virtual care in the future – this finding was consistently observed for all technologies. As per the technology acceptance model (TAM), the adoption of technology can be forecasted by perceived usefulness and perceived ease of use.^21^ It might be hypothesised that a more in-depth understanding of the risks associated with virtual care, as well as some potential digital fatigue by respondents with high digital literacy, may have a negative impact on their interested in future use. Other variables, such as social influence, facilitating conditions, trust, privacy, perceived risk, technological anxiety and resistance towards technology may also play an important role and could contribute to the variations observed.^22^

Finally, it is important to note that, among all dimensions of quality of care evaluated in this study, impact on healthcare equity was considered as least positive. These findings echo recent results by Chang JE (2021) that recognise important differences based on community characteristics in both the primary mode of remote care used and the types of barriers experienced.^23^ Our findings suggest that some patient groups may be particularly less engaged in the future use of remote care, including older persons. Reed et al (2020) observed that, for example, patients aged 65 years and over were less likely than patients aged 18 to 44 years to choose telemedicine.^24^ As mobile devices are used in most video visits, strategies to increase availability and access in these groups, as well as the co-design of user-centred solutions, may represent valuable opportunities to engage these patients.^25^ These differences in preferences by type of technology may indicate that access is not uniform.^24,26^ Despite the trend of a lower willingness in future use in older persons for most technologies, it is important to note that there appears to be nuance in choice of telephone visits and other types. For example, the proportion of participants wanting to keep using telephone consultations progressively increases with age. These results suggest important aspects of the impact of age and intended future use of different remote care choices and highlight that some remote care options may better meet specific patient groups’ needs than others.

## Strengths and limitations

A major strength of the study is the large sample size which gives more reliable results through greater statistical power, covering four different healthcare settings (UK, Germany, Italy, Sweden). The international scope of this work offers a wider lens in which to consider the perceived impact on quality, safety, and equity on primary care, and compare across a variety of settings. The sample populations were nationally representative, ensuring results accurately reflect the population in each national context. Furthermore, this study performed a detailed characterisation of the sample, including through the eHEALS health literacy tool.

The survey was conducted online meaning there is a risk of selection bias, as individuals with poor access to digital technology and the internet are less likely to have participated in the survey. As participants were only included if they were able to speak the resident country’s language, a proportion of the population were excluded from participation, likely including some vulnerable groups such as migrant workers and refugees. The non-randomised sample used in the study is a further limitation as its use makes the results less generalisable than the results of a randomised sample. However, this limitation was partly mitigated by ensuring the samples analysed were nationally representative. Thirdly, reporting bias is also widely recognized as a limitation of self-reported surveys assessing self-assessed behaviour.^27^ While the anonymity of the survey may have helped to reduce reporting bias, the possibility of this bias persisting should still be acknowledged as the study measured reported use rather than actual use.

## Implications for research and policy

This study is a starting point in considering the perspectives of the public across different health systems, as end-users of virtual care solutions in Primary Care through the COVID-19 pandemic and beyond. Discrepancies in experience rates between different groups of patients (particularly older persons or those with low digital health literacy), require further exploration to determine the risk of virtual care services amplifying existing inequities in healthcare. Creating effective digital health solutions is an opportunity that can be easily missed if digital design and implementation does not consider the needs and preferences of lower health literacy groups.^28^ There is, therefore, a need to develop digital health solutions that specifically meet the needs of lower health literate individuals, in order to increase overall acceptability and experience.^29^

Equity was the dimension that participants least often reported a positive impact - proactive efforts to address remote care barriers associated with specific groups of patients are therefore critical, to ensure the wide-scale implementation of virtual care does not entrench existing health disparities in health access in already underserved communities.^23^ After the emergency phase of the pandemic, we need to understand what worked well and for which patients, incorporating the lessons learned into new strategies to improve equity and digital inclusion.

Timeliness and efficiency were the most reported positive aspects of virtual care solutions, which offers the opportunity for further research to understand how these impact other domains of quality of care, particularly patient experience, and healthcare system savings. Until now, evidence on the cost-cutting potential of remote care has not been met with the same vigour as evidence-generation on their role in improving health outcomes.^30^

## Conclusion

The study provides new evidence on the topic of the perceived impact of using virtual care technologies in Primary Care during COVID-19, evaluating the use and perceptions on a range of virtual care tools. Future work must examine the drivers and determinants of positive experiences using remote care to co-create a supportive environment that ensures equitable adoption and use across different patient groups. Comparative analysis between countries and health systems offers the opportunity for policymakers to learn from best practices internationally.

## Data Availability

Data can be requested to the first author (ALN) upon reasonable request. Requests will be considered on an individual basis. Data sharing is not guaranteed.

