## Supplementary File 1 for "Use and impact of virtual primary care on quality and safety: the public’s perspectives during the COVID-19 pandemic"

**Online supplementary file**

**Supplementary Table 1. Perceived impact of telephone consultations on quality of care, by country**

| **Impact of telephone consultations** | **UK**  **N (%)** | **Germany**  **N (%)** | **Italy**  **N (%)** | **Sweden**  **N (%)** |
| --- | --- | --- | --- | --- |
| Positive | 715 (55.5) | 596 (54.3) | 480 (63.7) | 311 (53.4) |
| No impact | 372 (28.9) | 410 (37.3) | 199 (26.4) | 229 (39.3) |
| Negative impact | 201 (15.6) | 92 (8.4) | 75 (9.9) | 42 (7.2) |
| *Excluded (‘don’t know’ or missing values)* | *617* | *1063* | *377* | *433* |

**Supplementary Table 2. Perceived impact of video consultations on quality of care, by country**

| **Impact of video consultations** | **UK**  **N (%)** | **Germany**  **N (%)** | **Italy**  **N (%)** | **Sweden**  **N (%)** |
| --- | --- | --- | --- | --- |
| Positive | 342 (39.4) | 469 (47.4) | 243 (45.3) | 192 (44.4) |
| No impact | 406 (46.8) | 420 (42.4) | 244 (45.4) | 190 (44.0) |
| Negative impact | 120 (13.8) | 101 (10.2) | 50 (9.3) | 50 (11.6) |
| *Excluded (‘don’t know’ or missing values)* | *1151* | *1171* | *594* | *583* |

**Supplementary Table 3. Perceived impact of chat consultations on quality of care, by country**

| **Impact of chat consultations** | **UK**  **N (%)** | **Germany**  **N (%)** | **Italy**  **N (%)** | **Sweden**  **N (%)** |
| --- | --- | --- | --- | --- |
| Positive | 297 (33.7) | 380 (39.3) | 335 (52.3) | 165 (38.0) |
| No impact | 440 (49.9) | 470 (48.6) | 242 (37.8) | 218 (50.2) |
| Negative impact | 144 (16.3) | 118 (12.2) | 64 (10.0) | 51 (11.8) |
| *Excluded (‘don’t know’ or missing values)* | *730* | *1193* | *490* | *581* |

**Supplementary Table 4. Perceived impact of online triage on quality of care, by country**

| **Impact of online triage** | **UK**  **N (%)** | **Germany**  **N (%)** | **Italy**  **N (%)** | **Sweden**  **N (%)** |
| --- | --- | --- | --- | --- |
| Positive | 449 (44.1) | 420 (41.0) | 227 (41.6) | 198 (43.3) |
| No impact | 397 (39.0) | 435 (42.5) | 251 (46.0) | 216 (47.3) |
| Negative impact | 171 (16.8) | 169 (16.5) | 68 (12.5) | 43 (9.4) |
| *Excluded (‘don’t know’ or missing values)* | *1138* | *1137* | *585* | *558* |

**Supplementary Table 5. Perceived impact of remote monitoring on quality of care, by country**

| **Impact of remote monitoring** | **UK**  **N (%)** | **Germany**  **N (%)** | **Italy**  **N (%)** | **Sweden**  **N (%)** |
| --- | --- | --- | --- | --- |
| Positive | 246 (30.6) | 367 (39.1) | 260 (46.7) | 95 (27.9) |
| No impact | 446 (55.4) | 468 (49.8) | 232 (41.7) | 204 (59.8) |
| Negative impact | 113 (14.0) | 104 (11.1) | 65 (11.7) | 42 (12.3) |
| *Excluded (‘don’t know’ or missing values)* | *1002* | *1222* | *574* | *674* |

**Supplementary Table 6. Perceived impact of patient-initiated services on quality of care, by country**

| **Impact of patient-initiated services** | **UK**  **N (%)** | **Germany**  **N (%)** | **Italy**  **N (%)** | **Sweden**  **N (%)** |
| --- | --- | --- | --- | --- |
| Positive | 595 (54.2) | 683 (60.8) | 516 (69.6) | 377 (62.9) |
| No impact | 411 (37.4) | 368 (32.8) | 180 (24.3) | 192 (32.1) |
| Negative impact | 92 (8.4) | 72 (6.4) | 45 (6.1) | 30 (5.0) |
| *Excluded (‘don’t know’ or missing values)* | *921* | *1038* | *390* | *416* |

**Supplementary Table 7. Perceived impact of virtual care technologies on quality of care**

| **Total** | **Positive**  **N (%)** | **No impact**  **N (%)** | **Negative**  **N (%)** | *Don’t know*  *N* | *Missing values*  *N* |
| --- | --- | --- | --- | --- | --- |
| Patient-centredness | 1,919 (45.2) | 1,730 (40.8) | 592 (14.0) | *2,085 (33.0)* | *0 (0.0)* |
| Effectiveness | 1,802 (46.5) | 1,720 (44.4) | 350 (9.0) | *2,453 (38.8)* | *1 (0.0)* |
| Safety | 1,822 (45.5) | 1,701 (42.5) | 480 (12.0) | *2,324 (36.7)* | *0 (0.0)* |
| Efficiency | 2,401 (55.7) | 1,553 (36.0) | 359 (8.3) | *2,012 (31.8)* | *1 (0.0)* |
| Timeliness | 2,793 (60.2) | 1,320 (28.5) | 526 (11.3) | *1,686 (26.7)* | *1 (0.0)* |
| Equity | 1,726 (42.9) | 1,916 (47.6) | 381 (9.5) | *2,403 (36.4)* | *0 (0.0)* |

**Supplementary Table 8. Willingness of future use per digital health technology, by country**

|  | **UK (%)** | **Germany (%)** | **Italy (%)** | **Sweden (%)** | **Total (%)** |
| --- | --- | --- | --- | --- | --- |
| Telephone consultations | 1,226 (67.1) | 857 (44.5) | 550 (52.3) | 563 (61.3) | 3,195 (55.9) |
| Video consultations | 858 (47.0) | 481 (25.0) | 425 (40.4) | 350 (38.2) | 2,114 (37.0) |
| Chat consultations | 639 (35.0) | 349 (18.1) | 383 (36.4) | 347 (37.8) | 1,718 (30.0) |
| Online triage | 845 (46.3) | 511 (26.5) | 345 (32.8) | 304 (33.2) | 2,005 (35.1) |
| Patient-initiated digital services | 1,030 (56.4) | 891 (46.3) | 679 (64.4) | 606 (66.1) | 3,206 (56.1) |
| Secure messaging systems | 888 (48.7) | 543 (28.2) | 570 (54.2) | 479 (52.2) | 2,480 (43.4) |
| None of these | 267 (14.6) | 477 (24.8) | 50 (4.8) | 68 (7.4) | 862 (15.1) |
| *Excluded values*  *(“Don’t know”)* | *194* | *235* | *79* | *97* | *605* |

**Supplementary Table 9. Willingness of future use per digital health technology, by age**

| **All countries** | **18-24 (%)** | **25-34 (%)** | **35-44 (%)** | **45-54 (%)** | **55+ (%)** | **X2** | **P-value** |
| --- | --- | --- | --- | --- | --- | --- | --- |
| Telephone consults | 234 (39.9) | 482 (47.6) | 471 (48.6) | 560 (51.4) | 1448 (54.3) | 46.513 | <.001 |
| Video consults | 191 (32.6) | 378 (37.3) | 383 (39.5) | 378 (34.7) | 784 (29.4) | 43.419 | <.001 |
| Chat consults | 193 (32.9) | 344 (34.0) | 323 (33.3) | 281 (25.8) | 576 (21.6) | 95.176 | <.001 |
| Online triage | 145 (24.7) | 355 (35.0) | 356 (36.7) | 371 (34.0) | 780 (29.2) | 39.752 | <.001 |
| Secure messaging | 206 (35.2) | 419 (41.4) | 398 (41.0) | 418 (38.3) | 1039 (38.9) | 7.856 | 0.097 |
| Patient-initiated services | 262 (44.7) | 507 (50.0) | 481 (49.6) | 568 (52.1) | 1389 (52.1) | 11.909 | 0.018 |
| None of these | 52 (8.9) | 90 (8.9) | 120 (12.4) | 142 (13.0) | 459 (17.2) | 61.091 | <.001 |
| Footnote. A total of 605 respondents reported ‘don’t know’ to this item and were excluded from this analysis. | | | | | | | |

**Supplementary Table 10. Willingness of future use, by gender**

| **All countries** | **Male (%)** | **Female (%)** | **X2** | **P-value** |
| --- | --- | --- | --- | --- |
| Telephone consults | 1500 (48.6) | 1695 (52.3) | 8.545 | 0.003 |
| Video consults | 1124 (36.4) | 991 (30.6) | 24.450 | <.001 |
| Chat consults | 842 (27.3) | 875 (27.0) | 0.074 | 0.786 |
| Online triage | 1011 (32.8) | 995 (30.7) | 3.131 | 0.077 |
| Secure messaging | 1175 (38.1) | 1306 (40.3) | 3.199 | 0.074 |
| Patient-initiated services | 1493 (48.4) | 1714 (52.9) | 12.654 | <.001 |
| None of these | 453 (14.7) | 409 (12.6) | 5.723 | 0.017 |
| Footnote. A total of 605 respondents reported ‘don’t know’ to this item and were excluded from this analysis. | | | | |

**Supplementary Table 11. Willingness of future use, by digital health literacy level**

| **All countries** | eHEALS <25 (%) | eHEALS ≥ 26 (%) | X2 | P-value |
| --- | --- | --- | --- | --- |
| Telephone consults | 2866 (52.1) | 329 (39.7) | 44.220 | <.001 |
| Video consults | 1962 (35.7) | 153 (18.5) | 95.742 | <.001 |
| Chat consults | 1588 (28.9) | 129 (15.6) | 64.405 | <.001 |
| Online triage | 1851 (33.7) | 155 (18.7) | 74.190 | <.001 |
| Secure messaging | 2275 (41.4) | 206 (24.9) | 81.183 | <.001 |
| Patient-initiated services | 2885 (52.5) | 321 (38.8) | 54.080 | <.001 |
| None of these | 660 (12.0) | 202 (24.4) | 93.889 | <.001 |
| Footnote. A total of 605 respondents reported ‘don’t know’ to this item and were excluded from this analysis. | | | | |

**Supplementary Table 12. Willingness of future use, by self-reported ethnicity**

| **All countries** | White (%) | Mixed / Multiple (%) | Asian (%) | Black / African / Caribbean (%) | X2 | P-value |
| --- | --- | --- | --- | --- | --- | --- |
| Telephone consults | 2192 (57.7) | 74 (46.8) | 29 (34.5) | 6 (18.8) | 43.287 | <.001 |
| Video consults | 1504 (39.6) | 62 (39.2) | 31 (36.9) | 6 (18.8) | 5.992 | 0.112 |
| Chat consults | 1260 (33.2) | 52 (33.1) | 22 (26.2) | 8 (25.0) | 2.741 | 0.433 |
| Online triage | 1399 (36.8) | 45 (28.5) | 21 (25.0) | 3 (9.4) | 19.267 | <.001 |
| Secure messaging | 1805 (47.5) | 72 (45.6) | 20 (23.8) | 9 (28.1) | 22.213 | <.001 |
| Patient-initiated services | 2147 (56.5) | 83 (52.5) | 53 (36.9) | 11 (33.3) | 20.380 | <.001 |
| None of these | 356 (9.4) | 12 (7.60 | 12 (14.3) | 3 (9.1) | 2.966 | 0.397 |
| 1. A total of 605 respondents reported ‘don’t know’ to this item and were excluded from this analysis.  2. Respondents reporting ‘Other’ for ethnicity were excluded from this analysis (n=55).  3. Respondents from Germany excluded from this analysis as all responses in this country were skipped due to GDPR reasons (n=2,199). | | | | | | |
